# Teletherapy for children with developmental disorders during the COVID-19 pandemic in the Philippines: a mixed-methods evaluation from the perspectives of parents and therapists

**DOI:** 10.1101/2021.05.04.21256662

**Authors:** Kathlynne F. Eguia, Catherine M. Capio

## Abstract

**Objectives:** As a response to the lockdown associated with COVID-19 in the Philippines, therapy services for children with developmental disorders shifted to telehealth (i.e., teletherapy). This study evaluated the delivery of teletherapy from the perspectives of parents and therapists.

**Methods:** Participants consisted of parents (n = 47) and therapists (n = 102) of children with developmental disorders who were receiving teletherapy during the lockdown. A mixed-methods triangulation design-convergence model was adopted; participants were invited to respond to an online survey with closed- and open-ended questions. Quantitative data were analyzed using descriptive and non-parametric inferential tests, while qualitative data were examined using thematic analysis.

**Results:** Overall satisfaction with teletherapy was positive, with parents reporting significantly higher satisfaction compared to therapists. Satisfaction was positively associated with the frequency of teletherapy sessions for parents, and with their years of experience for therapists. The top enabling factors were family participation and effective communication. The main challenges were time constraints and difficulty with instruction and monitoring associated with the two-dimensional nature of teletherapy. The benefits included parents’ empowerment and enhanced understanding of their children’s needs.

**Discussion:** The shift to teletherapy facilitated a heightened focus on family-centered care. The evaluation findings suggest that the general satisfaction with teletherapy and the benefits associated with family-centered care will likely promote teletherapy as a service delivery mode to continue beyond the pandemic.

## Introduction

Following decades of rapid evolution of the internet and digital technologies, telehealth had been explored for patients who are physically located where health services may be difficult to obtain – such as rural areas or less developed territories (Farmer & Muhlenbruck, 2001). As lockdowns were implemented in many regions to contain the spread of COVID-19, telehealth became a critical component of health service delivery, including the provision of therapy for children with developmental disorders. Developmental disorders start in early childhood, and are characterized by impairments in physical, social, academic or occupational functions which require long-term management involving multiple specialists (Stuckey & Domingues-Montanari, 2017; Valentine et al., 2020). A direct impact of a COVID-19 lockdown to a child with developmental disorders is disruption of therapy, compromising the wellbeing of not only the child, but of the whole family.

The COVID-19 lockdown in the Philippines began in March 2020 and has been evolving as the severity of community transmission fluctuates. Therapy centers for children with developmental disorders closed when the lockdown began, and specialists shifted service delivery to telehealth (i.e., teletherapy). Such shift is advocated as a pediatric therapy response to the pandemic (Rao, 2021), with video consultations being the most important service delivery strategy (Alsem, Berkhout, & Buizer, 2020). We suggest that foundational to the shift to teletherapy is a family-centered care approach, which promotes family involvement in setting goals to improve clinical outcomes and family wellbeing (Franck & O’Brien, 2019). Studies in Western contexts have shown advantages with family involvement, such as parent-mediated telehealth being generally feasible, acceptable and beneficial for parents of children with developmental disorders (Pickard et al., 2016).

While we have positive expectations of teletherapy, it is important to understand its implementation in local contexts. In the Philippines, therapists typically deliver services in person and teletherapy was not commonly utilized prior to the pandemic. Parents, on the other hand, typically do not handle therapy activities for their children, to the extent that teletherapy would likely require. An evaluation is therefore important to identify the factors that could influence teletherapy service delivery (Pickard et al., 2016). Such evaluation could enable improvements to help mitigate the negative effects of a lockdown on the child’s development and the family’s wellbeing. This study, therefore, evaluated teletherapy delivery for children with developmental disorders in the Philippines during the COVID-19 pandemic, from the perspectives of therapists and parents. Specifically, (1) teletherapy service provision was characterized, (2) satisfaction and confidence were rated, and (3) enabling and challenging factors were explored.

## Materials and Methods

### Participants

Two groups of participants were recruited through convenience sampling: parents (n = 47) and therapists (n = 102) of children with developmental disorders who were receiving teletherapy during the lockdown in the Philippines. The therapists consisted of occupational therapists, physical therapists, speech-language therapists, and special education teachers. Recruitment was conducted through the social media pages of therapy centers and advocacy groups.

### Design and Procedures

The study was approved by the research ethics committee of the corresponding author’s affiliation (Reference: 2019-2020-0448). Participants had opportunities to seek clarifications, provided informed consent, and participated anonymously. A mixed-methods triangulation design-convergence model (QUAN + QUAL) was adopted to achieve well-substantiated evaluation conclusions (Creswell & Plano Clark, 2018). Participants were invited to respond to online surveys which included close-ended and open-ended questions. Details of service provision (i.e., digital platforms used, diagnosis of children, frequency of sessions) were reported through close-ended questions. Satisfaction and confidence with teletherapy were rated using five-point Likert-type scales (1 = very low, 5 = very high), in terms of communication, technology, and overall delivery. Parents also rated their satisfaction with their child’s participation and their confidence with family participation. Therapists rated their confidence with their strategies and service delivery. Factors that facilitated (enablers) and hindered (challenges) the conduct of teletherapy were examined through open-ended questions (e.g., What factors make it easier for you to participate during your child’s teletherapy?). Data were gathered from September to October 2020.

### Data analysis

Quantitative data from the close-ended questions were analyzed using descriptive statistics; non-parametric inferential statistics were performed because the ratings were not normally distributed (Shapiro-Wilk test, all *p* < 0.01). Responses to open-ended questions were examined through an inductive and semantic approach to thematic analysis, guided by a realist framework (Terry et al., 2017) and using the six-phase analytic process recommended by Braun and Clarke (2021). The six phases are: (1) familiarizing with the data, (2) generating codes, (3) generating initial themes, (4) reviewing and developing themes, (5) refining, defining and naming themes, and (6) writing the analysis. SPSS 26.0 was used for statistical analysis and NVivo 12.0 supported thematic analysis.

## Results

### Service provision

Based on the combined responses of parents and therapists, the most commonly used digital platform for teletherapy provision was Zoom (72%), followed by Facebook Messenger (28%) and Viber (16%). The other freely available platforms that were identified include Google Meets (9%), Skype (2%), and FaceTime (2%). Participants were able to report more than one platform; hence the percentages do not sum up to 100%. The diagnoses of the children who were receiving teletherapy included autism, Down Syndrome, global developmental delay, cerebral palsy, attention deficit hyperactivity disorder, and learning disability.

Parent respondents reported that their children were participating in teletherapy with an average of 2.31 (*S*.*D*. 1.62) one-hour sessions per week. Prior to the pandemic, the children were attending face-to-face therapy with an average of 2.73 (*S*.*D*. 2.49) sessions per week. The reduction in frequency of weekly therapy was not significant (*t*(41) = -1.41, *p* = 0.17). The therapists were delivering an average of 9.56 (*S*.*D*. 8.19) teletherapy sessions per week. Their average face-to-face therapy sessions prior to the pandemic was 29.40 (*S*.*D*. 13.22), which shows a significant reduction (*t*(101) = -16.47, p < 0.001) of workload during the pandemic. The therapists’ mean years of professional experience was 5.81 (*S*.*D*. 5.38).

### Evaluation ratings

The mean and 95% confidence intervals of the participants’ evaluation ratings are summarized in Table 1. Satisfaction with teletherapy was significantly higher among parents compared to therapists in terms of communication during teletherapy (*U* = 514.50, *p* < 0.001), technology support (*U* = 782.00, *p* = 0.007), and overall service delivery (*U* = 745.00, *p* = 0.002).

**Table 1.** Satisfaction and confidence ratings of parents and therapists on components of teletherapy delivery.

| Criterion | Parents (n = 49)<br>Mean (95% C.I.) | Therapists (n = 102)<br>Mean (95% C.I.) |
| --- | --- | --- |
| Overall satisfaction | 4.00 (3.62 – 4.38) | 3.58 (3.44 – 3.73) |
| Communication during teletherapy | 4.60 (4.28 – 4.91) | 3.69 (3.54 – 3.84) |
| Technology | 4.28 (3.91 – 4.65) | 3.74 (3.59 – 3.90) |
| Child's participation | 3.60 (3.08 – 4.12) |  |
| Family's participation (confidence) | 3.84 (3.34 – 4.34) |  |
| Knowledge of teletherapy strategies (confidence) |  | 3.51 (3.37 – 3.66) |
| Delivery of teletherapy (confidence) |  | 3.50 (3.35 – 3.65) |

Note: Five-point Likert-type scale; higher ratings represent higher satisfaction/confidence.

Among the parents, the frequency of their children’s teletherapy sessions was positively associated with their overall satisfaction (*r* = 0.399, *p* = 0.048). For the therapists, the frequency of delivering teletherapy sessions was positively associated with the satisfaction with their communication (*r* = 0.214, *p* = 0.044). Therapists’ years of experience was also positively associated with their overall satisfaction (*r* = 0.262, *p* = 0.014), confidence with strategies (*r* = 0.233, *p* = 0.029), and confidence with delivery (*r* = 0.316, *p* = 0.003).

### Enablers

The themes that were identified from the qualitative data analysis are summarized in Table 2. For parents, the most frequently reported enabler of teletherapy was *effective communication* with the therapist which includes interactions of the therapists with the child and their family members. This theme refers to therapists’ actions that shared knowledge, delivered instructions to family members, responded to parents’ queries, and provided immediate feedback to both the child and the family members. Shared knowledge included those that related to the developmental disorder of the child, and the rationale for the planned activities. The second factor was *flexibility of the therapist*, which refers to anticipating the child’s needs, modifying activities in response to the child, and maintaining engagement despite changes in the child’s disposition. The final factor according to parents was *timely and effective preparation* prior to each session. This includes provision of a general orientation to teletherapy for parents, listing of required materials ahead of each session, and providing illustrations to aid preparation of the designated space and materials at home.

**Table 2.** Themes that represent the enablers, challenges, and future insights in relation to teletherapy from the perspectives of parents and therapists of children receiving teletherapy.

|  | Parents | Therapists |
| --- | --- | --- |
| Enablers<br>(factors that help the delivery of teletherapy) | • Effective communications with the therapist<br>• High level of flexibility by the therapist<br>• Timely and effective preparation prior to sessions | • Adequate family preparation and participation<br>• Reliable technology and developed technology literacy<br>• Adequate professional preparation and support<br>• Available physical/material resources |
| Challenges<br>(factors that make teletherapy challenging) | • Conflicts with parents' time due to competing demands<br>• Limitations in physical/material resources at home<br>• Lack of confidence with following instructions Difficulties with child's disposition for online medium | • Difficulties with instruction and monitoring<br>• Inconsistency and limitations of technology<br>• Heightened physical/mental demands<br>• Limitations in family understanding and participation |
| Future implementation<br>(factors that can be considered for future teletherapy) | • Practical benefits that relate to convenience and time<br>• Enhanced family understanding and accessibility | • Perceived benefits that relate to families and therapists<br>• Considerations for future teletherapy practice<br>• Perceived costs that relate mostly to therapists |

The most widely reported enabling factor by therapists was *family participation and preparation*. This refers to family members who were motivated, collaborative, and participative during teletherapy sessions. For instance, one therapist noted that “parent involvement helped facilitate the activities based on the directions provided online”. This theme also refers to coordination with parents to identify the available resources in the home environment and setting them up prior to each teletherapy session. The next important enabler for therapists was *reliable technology and literacy* to use said technology. While this theme mostly refers to stable internet connections, it also includes gadgets (e.g., multi-angle cameras, earphones) and the ability to present information through technology (e.g., digital boards, slides). Adequate *professional preparation and support* was an enabler, which took the form of webinars and online materials about telehealth (e.g., practice guidelines), and professional communities (e.g., chat groups). For instance, such groups were reported to have enabled therapists to “brainstorm and collaborate with colleagues who are also doing teletherapy”. The *availability of resources* was the final enabler for the therapists. Digital resources refer to online worksheets, games and platforms (e.g., Twinkl, Boom Learning Cards); material resources refer to toys, learning materials (e.g., activity boards, puzzles), and exercise equipment (e.g., gym ball); physical resources refer to designated teletherapy spaces in the homes of the participants.

### Challenges

The challenge most reported by parents was *time constraints related to competing demands*. During the lockdown, most parents reported having to work from home and scheduling conflicts were inevitable (e.g., online work meeting and teletherapy appointment). Other competing demands included household chores and unforeseen circumstances (e.g., accidents). *Limitations in resources* at home also presented a challenge, and these include limited toys and equipment and lack of indoor space designated for teletherapy. A *lack of confidence with implementing instructions* during teletherapy was also reported. This refers to a sense of uncertainty associated with minimal training and preparation, as parents noted that they “are not professionals and it is quite difficult to follow (up) with planned activities after each session”. Finally, the parents noted challenges with the *child’s disposition* in relation to the online platform as some children required more time to adjust “since they are inside their comfort zone and maybe do not feel the need to participate” in activities at home.

The most frequently reported challenge for therapists was the *difficulty with instruction and monitoring*. This theme is generally associated with the therapists’ inability to provide tactile prompts and non-verbal cues for the child. Without the ability to touch or hold the child, the therapists were faced with limited input (i.e., visual, auditory only) compared to face-to-face therapy sessions. This theme also related to the inability to demonstrate to parents the actual grading of assistance (i.e., typically sensed through tactile means) that they needed to provide to their child. In terms of monitoring the child’s response, therapists also noted that non-verbal cues from the child are easily missed which subsequently leads to inadequate responses by the therapist. Inconsistency and *limitations of technology* was also reported by therapists frequently. This factor was largely associated with unstable internet connection, but several references were found about inadequate gadgets which the therapists themselves had to purchase. Difficulties were also experienced with managing relatively unfamiliar equipment (e.g., multi-angle cameras) and software (e.g., Zoom).

Therapists also reported *heightened physical and mental demands*, which refer to increased time and energy requirements associated with preparing for teletherapy sessions. Preparation includes searching online materials (e.g., YouTube videos), learning to use online resources, and integrating them with structured activities. Fatigue and physical exhaustion were noted to exceed those experienced during face-to-face therapy. Mental health challenges were experienced in the form of anxiety, stress, and mental fatigue. The final challenge reported by therapists was the *limitation in understanding and participation by family* members. This refers to therapists’ perceptions of lack of motivation, heightened frustration, and inconsistent participation by family members during teletherapy. This theme is also associated with time constraints (e.g., parents are too busy) and inadequate coordination among family members (e.g., multiple caregivers that respond differently).

### Benefits and future implementation

The parents reported *practical benefits* from teletherapy which include access to services in the comfort and safety of their home during the pandemic, and time saved from not having to travel to the clinic. Parents also reported *enhanced understanding* of their child’s needs and how each member of the family has roles in addressing those needs. For instance, one parent noted that therapists were “not just coaching, but also teaching me how to deal with my child’s behaviors”, suggesting that participating in teletherapy improved the parents’ ability to respond to the child. It was also noted that teletherapy shifted the focus on the child’s activities and routines that involved the participation of not only the parents but of other members of the family (e.g., grandparents, siblings). There was also *greater accessibility*, particularly for parents who are based overseas and previously did not directly interact with their children’s therapists.

The therapists similarly *perceived benefits* associated with parents’ enhanced empowerment in engaging with their children in their natural environments. For instance, it was noted that “parents became more aware of their child’s needs after doing teletherapy… parent involvement with goal setting also improved.” Several therapists reported that some goals were, in fact, achieved faster during teletherapy compared to face-to-face therapy prior to the pandemic.

A number of therapists acknowledged that teletherapy may persist as a service delivery model beyond the pandemic, and considerations for *future implementation* were identified. A key theme that surfaced is that therapists need to set parents’ expectations at the initial stage of teletherapy. The distinction of teletherapy from face-to-face sessions should be made early and clearly. Therapists engaging in teletherapy also need to be more deliberate with parent coaching strategies, and close collaboration is needed during planning and implementation of activities. Finally, the therapists acknowledged that practice standards need to be developed to support future implementation.

## Discussion

This study evaluated the delivery of teletherapy for children with developmental disorders in the Philippines during the COVID-19 lockdown, and identified factors to facilitate improved service delivery. While the shift to telehealth was sudden, it appears that for those who were able to access teletherapy, the needs of the children were met as evident in the relatively stable frequency when comparing face-to-face and teletherapy sessions as reported by parents. The wide range of freely available online platforms seems to have provided service delivery options, but the preference for the use of Zoom is consistent with those observed in the education and business sector during the pandemic (Mahr, Cichon, Mateo, Grajeda & Baggili, 2021). Zoom is known to have a flexible bandwidth requirement which automatically changes to suit the capacity of the available internet connection (e.g., 3G, WiFi) (Zoom Help Center, 2021). In the context of a developing country such as the Philippines, where internet speed continues to lag (Mingas, 2020), this flexibility of Zoom makes it a suitable platform for telehealth. Despite this, therapists identified unstable internet connection as one of the main challenges in delivering teletherapy. A further issue related to technology is the availability of devices, which had been raised as a concern where families who do not have access to laptops or tablets are potentially excluded from access to telehealth services (Alsem, et al., 2020). These findings suggest that a wider implementation of teletherapy in the Philippines could be hindered by current technology limitations which are mostly linked to economic resources.

Parents and therapists were generally satisfied with teletherapy service delivery, but more so the parents. Parents who participated in more frequent sessions tended to have higher overall satisfaction. Presumably, greater exposure and participation with teletherapy facilitated effective communication, which is the most widely reported enabler by parents. Therapists’ relatively lower overall satisfaction is likely linked to the difficulties with instruction and monitoring, as a consequence of being limited to two-dimensional interactions (i.e., visual, audio). However, those who have longer professional experience tended to have better overall satisfaction and confidence. This highlights the role of professional development and expertise despite teletherapy being a novel service delivery model for therapists in the Philippines. Indeed, professionals shifting to telehealth have been shown to draw from their existing professional knowledge and skills to educate and assess their patients in the online service model (Heckemann, Wolf, Ali, Sonntag, & Ekman, 2016). Therapists who delivered teletherapy more frequently also tended to be more satisfied with their communication, further emphasizing the benefits associated with professional experience.

### Highlighting family-centered care

The commonly reported enablers highlight that effective teletherapy is built on family-centered care, which is based upon a partnership of healthcare professionals with family members, through which the needs of the child are met (Kokorelias et al., 2019). For the parents, effective communication with therapists is the most important enabler, which is logically at the center of their partnership. This converges with the therapists’ perspective, where the most important enabler was family participation. The reported involvement of parents across goal-setting, activity planning, physical set-up, and implementation of activities is a demonstration of family-centered care being a systematic creation of a functioning partnership (Dunst, Trivette, & Hamby, 2007). As the pandemic raged, the main driver for teletherapy was the need to resume services in the face of a lockdown. While it was not deliberate, the opportunity for therapists to interact with family members in the home context allowed a shift to family-centered care, yielding a host of benefits.

Both parents and therapists recognized that a key benefit of teletherapy was the enhanced understanding and empowerment for parents. Indeed, it has been shown that empowerment of parents is a critical aspect in family-centered care and is associated with access to support from therapists (Fordham, Gibson & Bowes, 2012). Empowered parents become capable of creating opportunities for active participation by their children in their natural environment (Dalmau et al., 2017), and are able to manage their family life better, support the child’s needs, and therefore achieve significant progress for the child and the rest of the family (Marshall, Coulter, Gorski, & Ewing, 2016; McWilliam, 2016; Pereira & Oliveira, 2017). Such benefits are evident in this current evaluation, as therapists noted that in some children, progress was faster during teletherapy compared to that prior to the pandemic.

It is logical to argue that a focus on family-centered care should be sustained beyond the pandemic, such that associated benefits will persist. Moreover, parents’ recognition of the benefits of teletherapy suggests that there will be a continuing demand for this service delivery mode even when face-to-face sessions are able to resume. Our evaluation findings show that the therapists are aware that they will need to be more deliberate with coaching the parents. Hence, we suggest that professional development support is needed, such that therapists might be able to reinforce their relationships with parents and with other professionals (Gracia et al., 2020).

### Wellbeing issues

Parents and therapists alike grappled with issues that undermined their wellbeing. These issues seem to be related to the changes that forced people to work from home. Changes in work patterns ultimately led to increased time and energy requirements. Parents talked about not being professionals, and therefore not having adequate knowledge to confidently implement the activities during teletherapy. This suggests that they perceived increased expectations on their roles, which had been associated with feelings of anxiety and insecurity in the context of telehealth (Longo, de Campos, & Schiariti, 2020). In this current context, it appears that parents’ anxiety was mitigated by the flexibility and accessibility of therapists.

Therapists, however, talked about fatigue despite a significant decrease in the volume of weekly therapy sessions during the pandemic. One specific source of fatigue was the increased time and effort required to prepare ahead of each session. For example, a therapist wanted “more clients but at the same time, taking in too much meant having a hard time preparing all necessary materials”. Another significant stressor is associated with managing the difficulties of two-dimensional interactions, where therapists could not depend on tactile and non-verbal functions. Moreover, therapists needed to make their tacit knowledge (i.e., professional knowledge which reflect practical and technical expertise (Edwards, Jones, Carr, Braunack-Mayer, & Jensen, 2004)) be explicit as they had to be verbalized and explained to parents. For experienced therapists, such process loads on their cognitive resources, presumably contributing to mental fatigue.

We acknowledge that the upheaval caused by the pandemic in the daily lives of people is taking a toll on everyone’s mental health (Nielsen & Levkovich, 2020). On top of this, our evaluation suggests that shifting to teletherapy as a response to a lockdown came with costs that impact the wellbeing of both parents and therapists. As the pandemic continues, teletherapy may be more sustainable if supportive mechanisms for the wellbeing of parents and therapists will be put in place.

### Limitations

As an evaluation that aimed to inform service delivery improvement, we believe that the findings presented here are relevant and important. However, we note that the participants were recruited through convenience sampling, and therefore are not representative of the populations of parents and therapists of children with developmental disorders in the Philippines. Moreover, because this evaluation was focused on examining the enablers and challenges to service delivery, the experiences of families who had not been able to access teletherapy was not explored. This is another area that needs to be examined to address issues of accessibility. The severity of COVID-19 community transmission in the Philippines continued to fluctuate following the data gathering period. It is possible that parents’ and therapists’ wellbeing have changed further in the more recent lockdowns.

### Conclusions

This evaluation showed that stakeholders viewed teletherapy as a satisfactory service model, which will potentially be in demand beyond the pandemic. It appears that in responding to the therapy needs of children with developmental disorders during the lockdown, a shift to family-centered care was critical. Moving forward, systematic efforts are needed to better prepare parents and therapists for a more deliberate family-centered care approach to sustain the apparent benefits gained from teletherapy. Finally, supportive programs are needed to mitigate the negative impacts of teletherapy service model on stakeholders’ wellbeing.

## Data Availability

The data that support the findings of this study are available from the corresponding author, CMC, upon reasonable request.

